# Influence of physical activity and sleep duration on the retinal and choroidal structure in diabetic patients: An SS-OCT study

**DOI:** 10.1101/2020.02.18.20024752

**Authors:** Sen Liu, Wei Wang, Zihan Qiu, Miao He, Wenyong Huang

## Abstract

**Purpose:** To assess the association between physical activity, sleep duration, sitting time, and alterations of posterior segment structures with swept-source optical coherence tomography (SS-OCT).

**Methods:** Patients with diabetic retinopathy (DR) were recruited, and diabetic patients without retinopathy (non-DR) who matched for age and duration of diabetes were used as control. The physical activity, siting time, and sleep duration were obtained by using standardized questionnaire. OCT parameters included: retinal nerve fibre layer (RNFL) thickness, ganglion cell inner plexiform layer (GC-IPL) thickness, retinal thickness, and choroidal thickness (CT). Linear regression was conducted to analyse the association.

**Results:** Each group included 116 diabetic patients. Average macular CT was positively correlated with metabolic equivalents (MET) only in the DR group, independent of age, gender, and other potential confounding factors (β = 1.163, P = 0.006). Average macular CT was also positively correlated with sleep duration only in the non-DR group, independent of age, gender, and other potential confounding factors (β = 10.54, P = 0.031). No correlation was found between MET, sleep duration, and other OCT parameters. Sitting time was not significantly correlated with OCT parameters either.

**Conclusions:** Physical activity and sleep duration are both positively correlated with macular choroidal thickness; this suggests that more time in physical activity and sleep benefit the retina, while there was no association between sedentary time and OCT parameters. Further studies are warranted to clarify the underlying mechanisms and the role of physical activity and sleep in CT alterations and DR.

## INTRODUCTION

Diabetes mellitus (DM) and diabetic retinopathy (DR) have become major public health issues globally. There were 415 million adults with DM across the world in 2015, and the figure will reach 642 million by 2040.[1] Among DM patients, 35% will develop DR, which is the leading cause of blindness in the working-age population.[2] Many risk factors have been identified as being associated with DR, including the glycemia level, duration of DM, smoking, obesity, and hypertension.[2] Physical activity, sedentary time, and sleep duration have also been implicated in the development and progression of DR.[3,4] Optical coherence tomography (OCT) has been widely used for DR patients in clinical practice. The advance of OCT technology gives us an opportunity to obtain more detailed anatomical information of the posterior segment, including each layer of the retina and the choroid.[5]

Identifying related factors for OCT parameters is informative for clinical evaluation of normal variations and understanding the pathogenesis of DR. Previous studies mainly focus on the relationship between OCT parameters and traditional risk factors of diabetes, such as insulin resistance, poor glycaemic control, and renal function.[6-8] Population-based studies have shown that physical activity is related to retinal microcirculation, which has long been established as an independent risk factor of diabetes.[9-11] Emerging evidence suggests that sleep duration and sleep quality may possibly affect the incidence of diabetes and DR, as well as being associated with insulin resistance and poor glycaemic control.[12-14] However, little is known about the relationship of physical activity, sleep, and posterior segment structure, including the choroid and retina, in diabetic patients.

Considering the alterations in each layer of the retina and choroid in DM patients, it is reasonable to hypothesise that physical activity and sleep may also strongly impact OCT parameters. Therefore, the objective of this study was to investigate the association between physical activity, sleep, and retinal and choroidal parameters assessed with the latest swept-source optical coherence tomography (SS-OCT).

## METHODS

### Participants

This cross-sectional study was performed at Zhongshan Ophthalmic Centre (ZOC), Sun Yat-sen University, China. The protocol of the study was approved by the Institute Ethics Committee of ZOC. The study was performed according to the guidelines of the Helsinki Declaration. All participants signed written informed consent before entering. Subjects aged 35 or more who had type 2 diabetes with treatment-naïve eyes were recruited. The patients of DR were considered as case group (DR group), and the patients without DR were set as control group (NDR group). The diabetic patients with and without DR were matched for age and duration of DM. Exclusion criteria were as follows: (1) history of serious systemic diseases other than diabetes, such as serious cardiovascular and cerebrovascular diseases, malignant tumour, renal diseases, and chronic obstructive pulmonary disease; (2) history of surgery, such as thrombolysis therapy or kidney transplantation; (3) presence of cognitive impairment, mental illness, or inability to complete the questionnaire and examination; (4) history of glaucoma, age-related macular degeneration, vitreous-macular diseases other than DR, or amblyopia; (5) history of ocular surgical interventions, such as retina laser or intraocular injection, glaucoma surgery, cataract surgery, or laser myopia surgery; and (6) abnormal refractive media (moderate to severe cataract, corneal ulcer, pterygium, or corneal turbidity), poor fixation, and other causes resulting in poor quality of fundus or OCT images.

### Systemic and laboratory parameters

General information, including age, sex, duration of diabetes, other systemic chronic diseases, and risk factors, was collected via questionnaires. Height, weight, systolic blood pressure (SBP), and diastolic blood pressure (DBP) were measured by an experienced nurse. Mean blood pressure (mean BP) = 1/3 SBP + 2/3 DBP. The serum creatinine (Scr), haemoglobin A1c (HbA1c), total cholesterol (TC), triglycerides (TG), serum uric acid, and C-reactive protein were determined in standardized lab.

### Physical activity and sleep duration

The International Physical Activity Questionnaire (IPAQ) was used to assess the physical activity status of the participants. Metabolic equivalents (MET) were then calculated according to the principles of the IPAQ to quantify physical activity.[15] Sitting time, a proxy for sedentary time, was also included in the analysis. We used the Pittsburgh Sleep Quality Index (PSQI) to assess sleep quality over a 1-month time interval. The PSQI, a self-reporting questionnaire, is widely used in clinical research due to its well-established reliability and validity. We used the sleep duration item mainly for statistical analysis.[16]

### Ocular examination

Comprehensive ocular examinations were conducted for all participants. The anterior and posterior segments were evaluated by slit-lamp biomicroscopy and ophthalmoscopy. The evaluations of best corrected visual acuity (BCVA) and intraocular pressure (IOP) were regularly performed. Ocular biometric parameters were obtained using optical low-coherence reflectometry (Lenstar LS900; Haag-Streit AG, Koeniz, Switzerland). Auto refraction was measured by an autorefractor (KR8800; Topcon, Tokyo, Japan) after pupil dilation.

### SS-OCT imaging

The SS-OCT (DRI OCT-2 Triton; Topcon, Tokyo, Japan) instrument was used to obtain high-definition retina and choroid images. The macular region was scanned using a 6*6 mm volume scan protocol, which presents the thicknesses of each layer of retina and choroid in 9-ETDRS sectors. Segmentation of different layers on the OCT images and construction of topographic maps were automatically rendered by the software program. Segmentation was carefully inspected and confirmed by one OCT technician who was masked as to the refractive status of the participants, and manual correction was performed when the software misjudged the borderline of each layer. The peripapillary region was scanned using a 360 degree, 3.4-mm-diameter circle that was centred on the optic disc. Scans were centred using internal fixation. The macular region was scanned using a 3.14 circle. Parameters assessed by SS-OCT include average thickness of the macular retinal nerve fibre layer (mRNFL), GC-IPL thickness, retinal thickness, macular choroidal thickness (MCT), thickness of the peripapillary retinal nerve fibre layer (pRNFL), and peripapillary choroidal thickness (pCT).

### Statistical analyses

We incorporated data of the right eye only into analyses. The Kolmogorov-Smirnov test was carried out to verify normal distribution. When normality was confirmed, a t-test was conducted to evaluate inter-group differences of demographic, systemic, and ocular parameters. The Fisher’s exact test was used for categorical variables. Bivariate scatter plots were made to display the potential factors affecting OCT parameters. Variables with a P value < 0.10 in univariate analyses were included in the multivariate analysis. Univariate and multivariate linear regression analyses were used to assess the associations of MET, sitting time, sleep duration, and OCT parameters. We conducted crude linear regression in model 1. Potential confounding factors were then adjusted in multivariate regression analysis, with model 2 adjusting for age and sex, and model 3 for age, sex, gender, axial length, duration of diabetes, haemoglobin A1c (HbA1c), mean blood pressure, cholesterol, body mass index (BMI), intraocular pressure (IOP), best corrected visual acuity (BCVA, logMAR unit), and OCT imaging time. We also assessed the average MCT stratified by MET tertile in patients without retinopathy and stratified by different status of sleep duration in patients with retinopathy, using ANOVA. A P value of < 0.05 was considered statistically significant. All analyses were performed using Stata Version 14.0 (Stata Corporation, College Station, TX, USA).

## RESULTS

### Demographic and clinical characteristics

A total of 232 participants (116 in each group) who met inclusion/exclusion criteria were included in the final statistical analyses. The demographic and clinical features of the participants are shown in Table 1. The mean age was 64.2 ± 7.2 years, and almost half of them were male. The patients with and without DR did not differ in most demographic and clinical parameters, except for higher systolic blood pressure in DR patients (136.6 ± 19.5 mmHg versus 130.4 ± 16.0 mmHg, P = 0.009).

**Table 1.** Demographic and clinical characteristics of included diabetic patients.

| Characteristics | All | Diabetic retinopathy | Non diabetic retinopathy | P-value |
| --- | --- | --- | --- | --- |
| No. of subjects | 232 | 116 | 116 | - |
| Female, % | 123(53.02%) | 63(54.31%) | 60(51.72%) | 0.693 |
| Age, year | 64.2±7.2 | 64.1±7.3 | 64.2±7.1 | 0.935 |
| Duration of diabetes, year | 11.7±7.0 | 11.7±7.5 | 11.7±6.5 | 0.996 |
| HbA1c, % | 7.3±1.6 | 7.5±1.8 | 7.1±1.3 | 0.062 |
| Height, cm | 160.2±9.0 | 160.5±8.6 | 159.8±9.4 | 0.548 |
| Weight, kg | 62.6±11.2 | 63.7±11.8 | 61.4±10.5 | 0.134 |
| Mean blood pressure, mmHg | 180.1±22.5 | 182.9±24.3 | 177.3±20.3 | 0.060 |
| Total cholesterol, mmol/L | 5.0±1.1 | 4.9±1.0 | 5.0±1.2 | 0.516 |
| Serum creatinine, µmol/L | 76.2±25.5 | 78.3±28.6 | 74.0±21.8 | 0.199 |
| Triglycerides, mmol/L | 2.3±1.7 | 2.3±1.6 | 2.2±1.8 | 0.757 |
| Serum uric acid, µmol/L | 366.0±109.5 | 353.4±101.8 | 378.6±115.8 | 0.080 |
| C-reactive protein, mg/L | 2.7±4.5 | 2.9±4.6 | 2.5±4.4 | 0.585 |
| BCVA, logMAR | 0.2±0.1 | 0.2±0.1 | 0.2±0.2 | 0.788 |
| Intraocular pressure, mmHg | 16.0±2.6 | 16.3±2.7 | 15.8±2.5 | 0.099 |
| Central corneal thickness, µm | 548.1±32.6 | 549.2±33.8 | 547.0±31.5 | 0.611 |
| Axial length, mm | 23.4±1.1 | 23.4±1.0 | 23.4±1.1 | 0.809 |
| Anterior chamber depth, mm | 2.4±0.4 | 2.4±0.5 | 2.4±0.4 | 0.611 |
| MET, kcal/kg·h | 2391.4±1331.9 | 2460.8±1366.9 | 2321.9±1298.1 | 0.428 |
| Sitting time, minute/d | 357.3±125.9 | 345.8±124.4 | 368.8±126.8 | 0.164 |
| Sleep duration, hour/d | 6.1±1.3 | 6.1±1.2 | 6.1±1.4 | 0.956 |

Mean blood pressure=Diastolic blood pressure+1/3\*(Systolic blood pressure -Diastolic blood pressure); BCVA=best-corrected visual acuity;
MET=metabolic equivalents (kilocalories per kilograms per hour

### Physical activity, sitting time, and OCT parameters

Figure 1 shows that higher MET was associated with increased choroidal thickness, with average MCT 167.9 ± 9.9 μm for the first tertile, 198.3 ± 10.9 μm for the second tertile, and 214.5 ± 12.4 μm for the third tertile (P < 0.001). The results of the linear regression of MET with OCT parameters are summarized in Table 2. There was a positive correlation of average MCT and MET in the DR group (β **=** 1.561, 95% confidence interval (CI): 0. 653 to 2.469, P = 0.001). In further adjusting of other factors, MET remains significantly associated with average MCT (P = 0.005 in model 2, P = 0.006 in model 3). None of the other OCT parameters—mRNFL thickness, GC-IPL thickness, retinal thickness, pRNFL thickness, and pCT—were significantly related to MET. The MET was not related to any OCT parameters in NDR patients. Moreover, when we considered sitting time as another parameter to reflect physical activity, no significant correlations were found between it and all OCT parameters in both groups (Table 3).

**Table 2.**
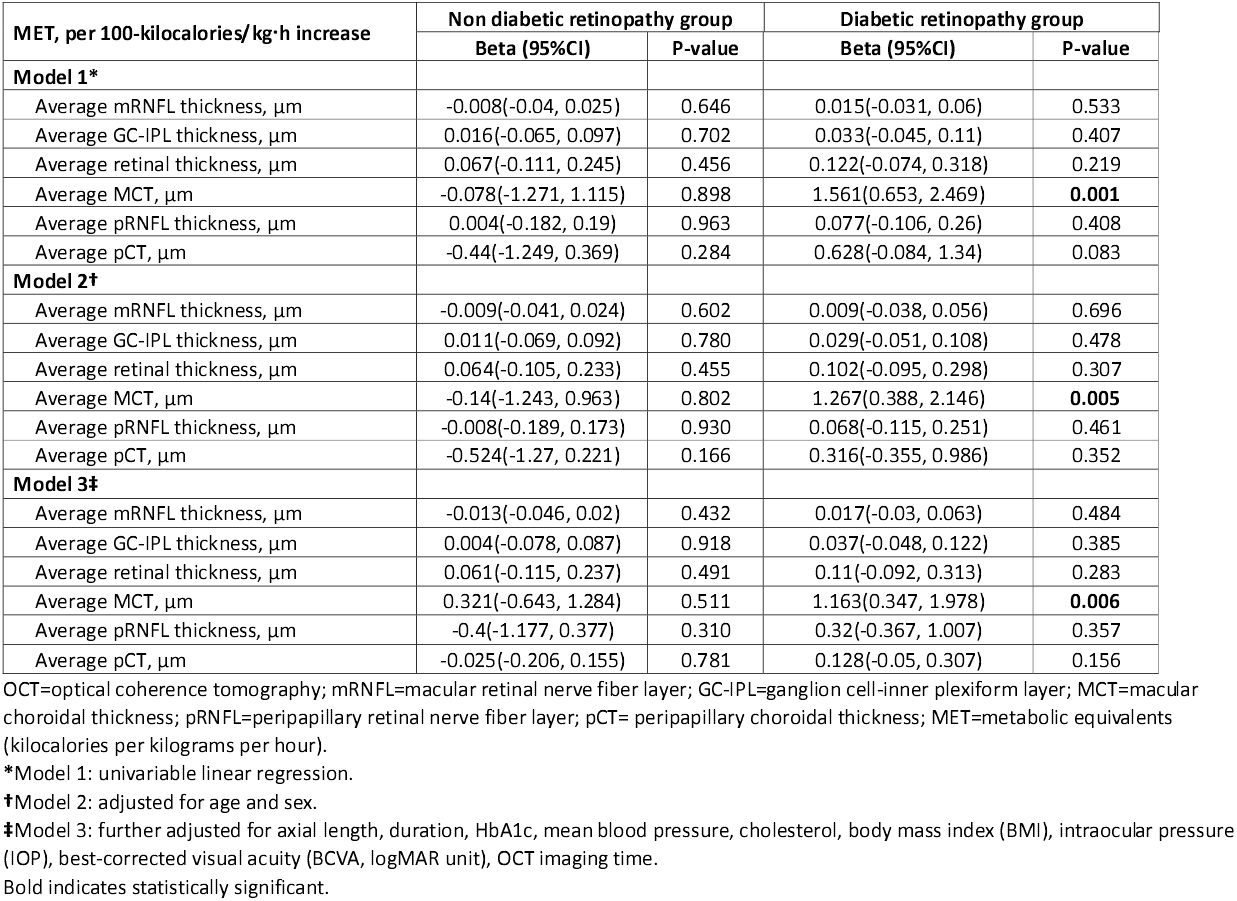
Relationship between physical activity and OCT parameters in both groups.

**Table 3.** Relationship between sitting time and OCT parameters in both groups.

| Sitting time, per 100-minutes increase | Non diabetic retinopathy group |  | Diabetic retinopathy group |  |
| --- | --- | --- | --- | --- |
|  | Beta (95%CI) | P-value | Beta (95%CI) | P-value |
| **Model 1\*** |  |  |  |  |
| Average mRNFL thickness, $\mu\text{m}$ | 0.248 (-0.078,0.575) | 0.135 | -0.052 (-0.556, 0.452) | 0.838 |
| Average GC-IPL thickness, $\mu\text{m}$ | -0.331 (-1.150,0.489) | 0.425 | -0.142 (-0.997, 0.713) | 0.743 |
| Average retinal thickness, $\mu\text{m}$ | 0.909 (-0.904,2.722) | 0.323 | -0.694 (-2.859, 1.471) | 0.527 |
| Average MCT, $\mu\text{m}$ | 7.191 (-4.946,19.328) | 0.243 | 1.199 (-9.267, 11.665) | 0.821 |
| Average pRNFL thickness, $\mu\text{m}$ | 0.659 (-1.236,2.554) | 0.492 | -0.160 (-2.226, 1.906) | 0.878 |
| Average pCT, $\mu\text{m}$ | 3.907 (-4.360,12.174) | 0.351 | -5.852 (-13.575, 1.872) | 0.136 |
| **Model 2†** |  |  |  |  |
| Average mRNFL thickness, $\mu\text{m}$ | 0.236 (-0.101,0.573) | 0.168 | -0.043 (-0.572, 0.487) | 0.874 |
| Average GC-IPL thickness, $\mu\text{m}$ | -0.336 (-1.172,0.500) | 0.428 | -0.138 (-1.041, 0.765) | 0.762 |
| Average retinal thickness, $\mu\text{m}$ | 0.165 (-1.612,1.941) | 0.855 | -1.523 (-3.737, 0.690) | 0.175 |
| Average MCT, $\mu\text{m}$ | 2.005 (-9.579,13.588) | 0.732 | -0.181 (-10.487, 10.125) | 0.972 |
| Average pRNFL thickness, $\mu\text{m}$ | 0.375 (-1.515,2.265) | 0.695 | 0.667 (-1.449, 2.783) | 0.533 |
| Average pCT, $\mu\text{m}$ | 0.666 (-7.195,8.527) | 0.867 | -5.799 (-13.205, 1.607) | 0.124 |
| **Model 3‡** |  |  |  |  |
| Average mRNFL thickness, $\mu\text{m}$ | 0.262 (-0.095,0.619) | 0.148 | 0.084 (-0.488, 0.655) | 0.772 |
| Average GC-IPL thickness, $\mu\text{m}$ | -0.404 (-1.313,0.505) | 0.380 | -0.558 (-1.599, 0.483) | 0.290 |
| Average retinal thickness, $\mu\text{m}$ | -0.002 (-1.985,1.980) | 0.998 | -1.953 (-4.420, 0.515) | 0.120 |
| Average MCT, $\mu\text{m}$ | -4.153 (-14.976,6.669) | 0.448 | -5.970 (-16.317, 4.376) | 0.255 |
| Average pRNFL thickness, $\mu\text{m}$ | -1.272 (-9.920,7.376) | 0.771 | -5.926 (-14.183, 2.330) | 0.157 |
| Average pCT, $\mu\text{m}$ | 0.040 (-1.961,2.040) | 0.969 | 0.656 (-1.598, 2.909) | 0.565 |

OCT=optical coherence tomography; mRNFL=macular retinal nerve fiber layer; GC-IPL=ganglion cell-inner plexiform layer; MCT=macular choroidal thickness; pRNFL=peripapillary retinal nerve fiber layer; pCT= peripapillary choroidal thickness; MET=metabolic equivalents (kilocalories per kilograms per hour).
\*Model 1: univariable linear regression.
†Model 2: adjusted for age and sex.
‡Model 3: further adjusted for axial length, duration, HbA1c, mean blood pressure, cholesterol, body mass index (BMI), intraocular pressure (IOP), best-corrected visual acuity (BCVA, logMAR unit), OCT imaging time.

**Figure 1.**
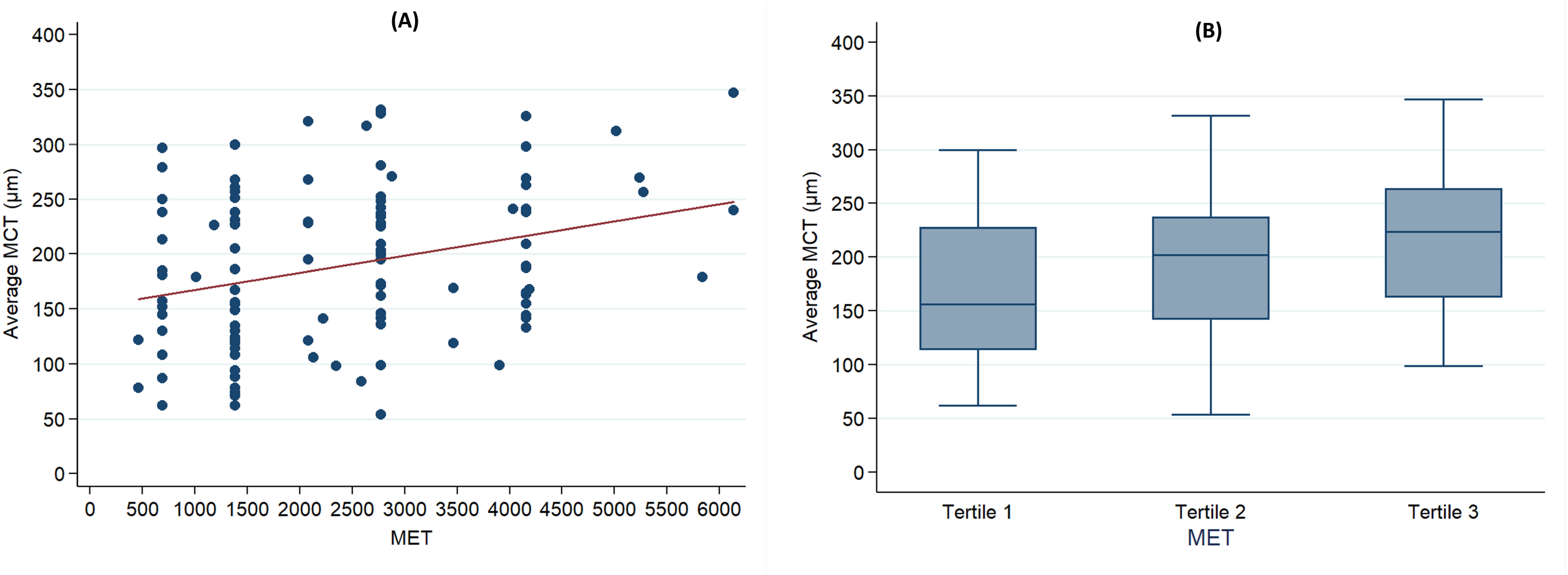
Choroidal thickness in macular region stratified by MET tertile in patients without retinopathy.

### Sleep duration and OCT parameters

For patients with NDR, average MCT was positively correlated with sleep duration (β = 16.02, 95% CI: 4.76 to 27.28, P = 0.006), as shown in Figure 2A. The average MCT was significantly different among the three subgroups (167.2 ± 20.0 μm versus 209.7 ± 9.6 μm versus 247.0 ± 21.3 μm, P < 0.001). The results of linear regression of sleep duration and OCT parameters are summarized in Table 4. After adjusting other factors, sleep duration remains independently correlated with average MCT in NDR patients, with β = 11.11 (95% CI: 0.03 to 22.19, P = 0.049) in model 2 and β = 10.54 (95% CI: 0.96 to 20.12, P = 0.031) in model 3. For OCT parameters in patients with DR, no significant association with sleep duration was noted (All P>0.05).

**Table 4.** Relationship between sleep duration and OCT parameters in both groups.

| Sleep duration, per 1-hour increase | Non diabetic retinopathy group |  | Diabetic retinopathy group |  |
| --- | --- | --- | --- | --- |
|  | Beta (95%CI) | P-value | Beta (95%CI) | P-value |
| **Model 1\*** |  |  |  |  |
| Average mRNFL thickness, $\mu\text{m}$ | 0.092 (-0.228, 0.412) | 0.568 | -0.432 (-0.958, 0.094) | 0.106 |
| Average GC-IPL thickness, $\mu\text{m}$ | -0.362 (-1.155, 0.432) | 0.368 | -0.221 (-1.117, 0.674) | 0.625 |
| Average retinal thickness, $\mu\text{m}$ | 1.282 (-0.471, 3.035) | 0.150 | -1.139 (-3.413, 1.135) | 0.323 |
| Average MCT, $\mu\text{m}$ | 16.020 (4.758, 27.282) | **0.006** | -3.915 (-15.360, 7.530) | 0.499 |
| Average pRNFL thickness, $\mu\text{m}$ | 0.983 (-0.915, 2.880) | 0.307 | -0.732 (-2.904, 1.439) | 0.505 |
| Average pCT, $\mu\text{m}$ | 6.174 (-1.688, 14.035) | 0.122 | -0.740 (-8.998, 7.519) | 0.859 |
| **Model 2†** |  |  |  |  |
| Average mRNFL thickness, $\mu\text{m}$ | 0.071 (-0.264, 0.407) | 0.674 | -0.398 (-0.926, 0.131) | 0.138 |
| Average GC-IPL thickness, $\mu\text{m}$ | -0.351 (-1.173, 0.472) | 0.399 | -0.192 (-1.098, 0.714) | 0.675 |
| Average retinal thickness, $\mu\text{m}$ | 0.462 (-1.288, 2.212) | 0.602 | -1.130 (-3.378, 1.117) | 0.321 |
| Average MCT, $\mu\text{m}$ | 11.110 (0.031, 22.188) | **0.049** | -2.611 (-13.491, 8.269) | 0.635 |
| Average pRNFL thickness, $\mu\text{m}$ | 0.938 (-0.934, 2.811) | 0.323 | -0.534 (-2.668, 1.601) | 0.621 |
| Average pCT, $\mu\text{m}$ | 3.134 (-4.357, 10.624) | 0.409 | 1.506 (-6.096, 9.108) | 0.695 |
| **Model 3‡** |  |  |  |  |
| Average mRNFL thickness, $\mu\text{m}$ | -0.031 (-0.363, 0.301) | 0.853 | -0.324 (-0.886, 0.237) | 0.254 |
| Average GC-IPL thickness, $\mu\text{m}$ | -0.313 (-1.153, 0.526) | 0.461 | -0.286 (-1.318, 0.746) | 0.583 |
| Average retinal thickness, $\mu\text{m}$ | 0.558 (-1.267, 2.384) | 0.545 | -0.548 (-3.029, 1.933) | 0.662 |
| Average MCT, $\mu\text{m}$ | 10.541 (0.962, 20.121) | **0.031** | -6.774 (-17.643, 4.094) | 0.219 |
| Average pRNFL thickness, $\mu\text{m}$ | 1.106 (-0.695, 2.908) | 0.226 | -0.181 (-2.436, 2.075) | 0.874 |
| Average pCT, $\mu\text{m}$ | 2.379 (-5.410, 10.167) | 0.546 | 2.136 (-6.060, 10.333) | 0.605 |

OCT=optical coherence tomography; mRNFL=macular retinal nerve fiber layer; GC-IPL=ganglion cell-inner plexiform layer; MCT=macular choroidal thickness; pRNFL=peripapillary retinal nerve fiber layer; pCT= peripapillary choroidal thickness; MET=metabolic equivalents (kilocalories per kilograms per hour).
\*Model 1: univariable linear regression.
†Model 2: adjusted for age and sex.
‡Model 3: further adjusted for axial length, duration, HbA1c, mean blood pressure, cholesterol, body mass index (BMI), intraocular pressure (IOP), best-corrected visual acuity (BCVA, logMAR unit), OCT imaging time.
Bold indicates statistically significant.

**Figure 2.**
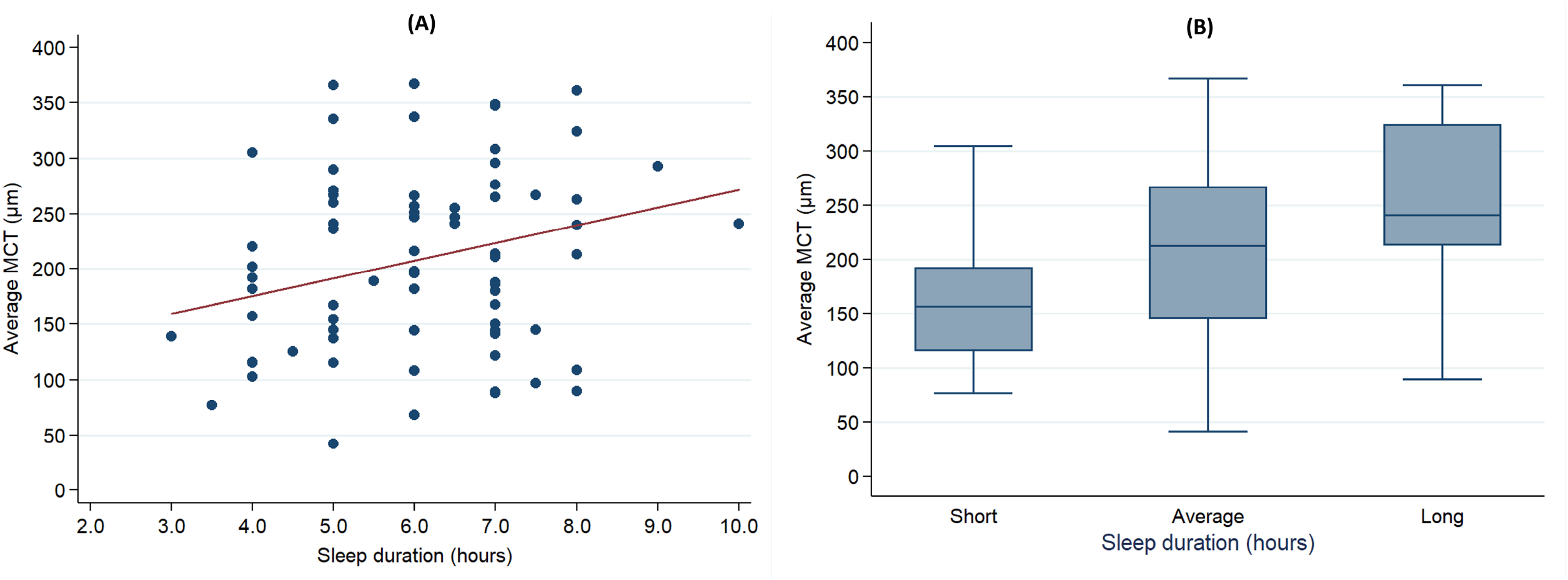
Choroidal thickness in macular region stratified by sleep duration in patients with retinopathy.

## DISCUSSION

The present study used SS-OCT to explore the association of physical activity and sleep duration with posterior segment structure in diabetic patients. The results suggested a thickened macular choroid in DR patients with more physical activity, but no association between sitting time and OCT parameters. We also found a positive correlation between sleep duration and the MCT in individuals without DR. To the best of our knowledge, this is the first study to correlate the OCT parameters centring on macula and optic nerve head with physical activity and sleep duration.

The choroid provides 85% of the blood supply to the eyeball and plays a very important role in normal retinal and choroidal functions. There are limited data on the relationship between physical activity and retino-choroidal microcirculation.[17-20] Our finding of an increased CT being associated with higher physical activity indirectly affirms the previous finding that a higher level of physical activity lowers the likelihood of DR due to altering the retinal microvascular function. Keel et al.[21] reported an inverse correlation between physical activity and retinal venular calibre in adolescents with type 1 diabetes, while Gopinath et al.[9] reported that higher levels of physical activity are associated with narrower retinal arterioles. Sayin et al.[22] reported that short-term dynamic exercise causes a significant increase in CT for at least 5 min following exercise. However, Hong et al.[23] did not find any changes of MCT in healthy subjects after 15 min on a treadmill. Alnawaiseh et al.[18] demonstrated that increased physical activity induced significant changes in optic nerve and macular perfusion. Nussbaumer et al.[24] reported that the effects of endurance exercise on retinal vessel diameters was age- and intensity-dependent. The findings of our study and previous studies, that physical activity may cause long-term effects on the choroid, may be dependent on varying disease status.

As to sedentary behaviour, we found it is not related to CT and other OCT parameters. Anuradha et al. reported that sedentary behaviour quantified by TV viewing time was significantly associated with the retinal microcirculation in an Australian population.[25] The Multi-Ethnic Study of Atherosclerosis (MESA) demonstrated that television viewing time is related to retinal vascular calibre only in non-Hispanic whites, but not in other ethnic groups.[10] The discrepancy of results in the aforementioned studies may be related to different definitions of sedentary behaviour and different ethnicities. The form of sedentary behaviour measured in the two studies mentioned above was TV viewing and playing computer/video games. This may cause confounding bias as time spent on electronic screens may affect retinal microcirculation independently. Consistent with the MESA study, we found no association of sedentary time with OCT parameters in this Chinese population.

Sleep deprivation may play a role in the pathogenesis of DR but remains unclear. Recent studies confirm that obstructive sleep apnoea syndrome (OSAS), characterized as sleep disturbance, is an independent risk factor for the presence and progression of DR.[12,14,26,27] Sleep duration has recently been reported to be related to DR risk, and it has been well documented that CT is changed in DR patients.[28,29] This study first assessed the relationship between posterior segment structures and sleep duration in diabetic patients, and found a positive correlation between MCT and sleep duration in diabetic patients without DR, but no significant association in DR patients. The relationship between sleep and CT needs further study.

This study suggests that more physical activity and more sleep are good for the retina and may reduce the risk of the onset of DR, but the underlying pathogenic mechanisms to explain these associations remain inconclusive. Hypoxemia, inflammation, oxidative stress, hormonal changes, insulin resistance, and disturbance of autoregulation in retinal microcirculation may possibly take part in the pathogenesis of choroid alteration leading to DR.[5,28,29]

The major strengths of the present study are: (1) the use of standardized questionnaires, such as the IPAQ and PSQI, to quantify the status of physical activity and sleep affords well-established reliability and validity of our data; (2) the use of SS-OCT enabled us to obtain non-invasive ‘optical slices’ of the posterior segment, including the retina and the choroid; and (3) the comprehensive consideration of potential confounding factors (age, gender, axial length, duration of diabetes, HbA1c, BP, cholesterol, BMI, IOP, BCVA, and OCT imaging time). This study also has certain limitations. First, we only analysed the most representative parameters of physical activity and sleep in the form of questionnaires, which may have recall bias. More objective assessment of more complex factors obtained from wearable devices may be a better method to reduce recall bias in the future.[30] Second, as this study utilized a cross-sectional design, we cannot infer causality from the observed associations. Further cohort studies are warranted to verify the findings of the present study.

In conclusion, the present study documented a significant relationship between physical activity, sleep duration, and retino-choroidal structures assessed by SS-OCT in Chinese patients with type 2 diabetes mellitus. Greater physical activity and longer sleep duration were correlated with thicker macular choroidal thickness in DR and NDR patients. Further studies are warranted to clarify the underlying mechanisms and the role of physical activity and sleep in CT alterations and DR progression.

## Data Availability

All dada were in the manuscript

## Acknowledgement

None.

## Statement of Ethics

All examinations involving human participants were performed in accordance with the ethical standards of ZOC ethical committee and the Helsinki declaration and its later amendments or comparable ethical standards. Informed consent was obtained from all participants.

## Author contributions

Design and conduct of the study (WW, YL, WH); Collection, management, analysis and interpretation of the data (ZQ, WW, YT, MH, LW, YL, WH); Preparation of the manuscript (ZQ, WW, WH); Review and final approval of the manuscript (all authors).

